# Combination of hotspot mutations with methylation and fragmentomic profiles to enhance Multi-Cancer Early Detection

**DOI:** 10.1101/2024.08.18.24312164

**Authors:** Thi Hue Hanh Nguyen, Hoang Giang Vu, Thi Tu Nguyen, Anh Tuan Nguyen, Vu Uyen Tran, Thi Luyen Vu, Thi Huong Giang Nguyen, Duy Nhat Nguyen, Trung Hieu Tran, Van Thien Chi Nguyen, Thanh Dat Nguyen, Trong Hieu Nguyen, Dac Ho Vo, Thi Tuong Vi Van, Thi Thanh Do, Minh Phong Le, Le Anh Khoa Huynh, Duy Sinh Nguyen, Hung Sang Tang, Hoai-Nghia Nguyen, Minh-Duy Phan, Hoa Giang, Lan N Tu, Le Son Tran

**Affiliations:** Medical Genetics Institute, Ho Chi Minh, Vietnam

**Keywords:** MCED, hotspot mutations, cfDNA, genetic and epigenetic alterations

## Abstract

**Background:** Multi-cancer early detection (MCED) through a single blood test significantly advances cancer diagnosis. However, most MCED tests rely on a single type of biomarkers, leading to limited sensitivity, particularly for early-stage cancers. We previously developed SPOT-MAS, a multimodal ctDNA-based assay analyzing methylation and fragmentomic profiles to detect five common cancers. Despite its potential, SPOT-MAS exhibited moderate sensitivities for early-stage cancers. This study investigated whether integrating hotspot mutations into SPOT-MAS could enhance its detection rates.

**Method:** A targeted amplicon sequencing approach was developed to profile 700 hotspot mutations in cell-free DNA and integrated into the SPOT-MAS assay, creating a single-blood draw workflow. This workflow, namely SPOT-MAS Plus was retrospectively validated in a cohort of 255 non-metastatic cancer patients (breast, colorectal, gastric, liver, and lung) and 304 healthy individuals.

**Results:** Hotspot mutations were detected in 131 of 255 (51.4%) cancer patients, with the highest rates in liver cancer (96.5%), followed by colorectal (59.3%) and lung cancer (53.7%). Lower detection rates were found for cancers with low tumor mutational burden, such as breast (31.3%) and gastric (41.9%) cancers. In contrast, SPOT-MAS demonstrated higher sensitivities for these cancers (51.6% for breast and 62.9% for gastric). The combination of hotspot mutations with SPOT-MAS predictions improved early-stage cancer detection, achieving an overall sensitivity of 78.5% at a specificity of 97.7%. Enhanced sensitivities were observed for colorectal (81.36%) and lung cancer (82.9%).

**Conclusion:** The integration of genetic and epigenetic alterations into a multimodal assay significantly enhances the early detection of various cancers. Further validation in larger cohorts is necessary to support broader clinical applications.

## INTRODUCTION

Multi-cancer early detection (MCED) utilizing circulating tumor DNA (ctDNA) in the bloodstream is a significant breakthrough in cancer screening. By employing a multimodal approach to capture various ctDNA molecular signatures, several MCED tests have demonstrated effectiveness in detecting multiple cancer types at early stages, thereby facilitating timely treatment and significantly improving patient outcomes. Liu *et al.* developed the Galleri test, an MCED assay designed to screen for over 50 types of early-stage cancers by detecting specific methylation patterns in ctDNA. In a retrospective study, the Galleri test achieved a specificity of 99.5%, an overall sensitivity of 51.5%, and a sensitivity of 76.3% for 12 particularly lethal cancers (1–3). The recent PATHFINDER prospective validation study further validated the clinical utility of the Galleri test, reporting a positive predictive value of 38% and a negative predictive value of 98.6% (2). These results highlight the promise of the ctDNA-based MCED method in clinical practice.

Despite the promising outcomes, ctDNA detection for early-stage cancers remains challenging due to its low abundance and molecular heterogeneity across different cancer types and histological subtypes (4). For instance, breast cancers are known to release lower concentrations of ctDNA compared to cancers with a higher mutational burden, such as lung and colorectal cancers (5). Notably, different breast cancer subtypes exhibit varying levels of ctDNA shedding; luminal subtypes tend to have lower ctDNA levels compared to HER2+ and triple-negative breast cancer (TNBC) subtypes (6, 7). Similarly, in non-small cell lung cancer (NSCLC), lung adenocarcinomas generally present with lower ctDNA levels, while centrally located squamous cell carcinomas often show higher levels of ctDNA. These differences underscore the complexity of ctDNA dynamics across cancer types and highlight the necessity of a multimodal approach that captures a broad spectrum of molecular signatures to enhance ctDNA detection and improve the performance of MCED assays.

We previously developed a multimodal assay, Screening for the Presence Of Tumor by Methylation And Size (SPOT-MAS), which integrates multiple ctDNA signatures, including methylomics and fragmentomics, to detect the five most common cancers in Vietnam: liver, breast, colorectal, gastric, and lung. While SPOT-MAS has demonstrated promising results in both retrospective and prospective validations, its detection sensitivities varied across different cancer types and stages, with the lowest rates observed for breast cancer (49.3%) and early-stage tumors (62.3% to 73.9% for stages I and II) (8). Tumor-derived genetic variants, along with changes in methylation, play a significant role in driving carcinogenesis in certain cancer types and have been leveraged in numerous studies to detect ctDNA in plasma (9–12). One prominent example is the CancerSeek test, developed by Ludwig Cancer Research at Johns Hopkins University, which employed a panel of mutations at 2,001 locations across 16 cancer-associated genes (*TP53, GNAS, PPP2R1A, HRAS, KRAS, AKT1, PTEN, FGFR2, CDKN2A, BRAF, EGFR, APC, FBXW7, PIK3CA, CTNNB1,* and *NRAS*) to detect eight cancer types (13).

It is thought that genetic mutations and epigenetic changes can occur either independently or concurrently, with potential bidirectional interactions during tumorigenesis (14, 15). To enhance the capacity of the SPOT-MAS assay in capturing ctDNA signals across multiple cancer types, we aimed to integrate the detection of hotspot genetic variants into its existing workflow focusing on methylation and fragmentomic features. However, using hotspot mutations for multi-cancer early detection poses certain challenges, such as the prior knowledge of recurrent mutations and the confounding effects of mutations linked to clonal hematopoiesis of indeterminate potential (CHIP) (12, 16). To address these challenges, we developed an in-house panel comprising 700 hotspot mutations selected from the Catalogue of Somatic Mutations in Cancer (COSMIC) database and somatic mutations identified in a large cohort of 1,100 Vietnamese cancer patients. We then evaluated the potential utility of this panel for detecting five common cancer types, either as a standalone approach or in combination with our SPOT-MAS assay (**Figure 1**).

**Figure 1.**
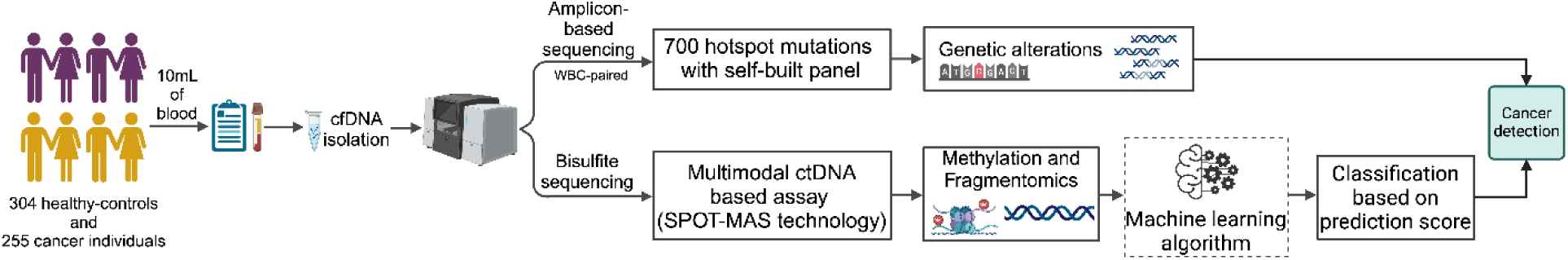
Diagram illustrates study design. Our study recruited 304 healthy controls and 255 cancer participants. From each participant, 10 ml of blood was drawn. Cell-free DNA (cfDNA) and genomic DNA (gDNA) were isolated for targeted sequencing using a 700-hotspot mutation panel. Mutation calling was performed, with CHIP mutations filtered out to confirm positive mutations. Another fraction of cfDNA was subjected to the SPOT-MAS assay, which uses bisulfite sequencing to profile methylomic and fragmentomic features for classification analysis by machine learning algorithms. The detection abilities of the hotspot mutation-based approach and the SPOT-MAS-based approach were evaluated individually and in combination.

## MATERIAL and METHODS

### Patients and sample collection

Blood samples were collected from 304 healthy controls and 255 cancer patients diagnosed with one of five cancer types: breast, colorectal, gastric, liver, and lung (**Table S1**). Cancer diagnoses were confirmed through imaging and/or histological analysis, depending on the type of cancer. Patients with prior treatment or metastasis were excluded from the study.

All participants provided written informed consent for their involvement and for the anonymization of their samples, clinical, and genomic data. The data were de-identified prior to analysis of the cohort.

### Cell-free DNA and genomic DNA sample preparation

Blood samples (10 mL) were subjected to a two-step centrifugation process (1,600×g for 10 minutes at 4°C, followed by 16,000×g for 10 minutes at 4°C) to separate plasma from cellular components. The buffy coat was carefully collected by gently pipetting the buff-colored layer, ensuring that the other blood components remained undisturbed. Both the plasma and buffy coat were then stored at −80°C.

Cell-free DNA (cfDNA) was extracted from the plasma using the MagMAX Cell-free DNA Isolation kit (Thermo Fisher, USA) on the KingFisher Flex Magnetic 96DW automated system, according to the manufacturer’s instructions.

Genomic DNA (gDNA) was isolated from the buffy coat using the GeneJET Whole Blood Genomic DNA Purification Mini Kit (Thermo Fisher, USA) following the manufacturer’s instructions. The isolated cfDNA and gDNA were recovered and stored in DNA LoBind tubes (Eppendorf AG) at −20°C if not used immediately, and the DNA concentration was measured using the QuantiFluor dsDNA system (Promega, USA).

### Amplicon-based sequencing

We profiled a panel of 700 hotspot mutations selected from the COSMIC database and somatic mutations identified in a cohort of 1,100 Vietnamese cancer patients suffering from breast, colorectal, gastric, liver, and lung cancer. Hotspot mutations were selected based on the following criteria: (1) reported hotspot mutations in the COSMIC database that are recurrent in the five cancer types, (2) reported actionable mutations, and (3) mutations found in tumor tissue from the Vietnamese cancer patients according to five types of cancers (breast, colorectal, gastric, liver and lung cancer). Compatible primer pairs were designed using Primer3Plus software (17) and synthesized by PhuSa Biochem (Ho Chi Minh City, Vietnam). Details of panel 700 hotspot mutations in 23 genes are listed in **Table S2**.

For multiplex PCR (mPCR), 3.2 ng of cfDNA was used to amplify specific DNA segments containing the targeted mutations. The mPCR reaction included 5 µL of primer mix at 0.5 µM and 25 µL of KAPA HiFi DNA Polymerase mastermix (Roche Sequencing Solutions, Indianapolis, IN, USA). The target enrichment thermocycler program consisted of denaturation at 98°C for 45 s, amplification with 25 cycles at (98°C for 20 s, 64°C for 1 min, 72°C for 5 min), final extension at 72°C for 5 min, and hold at 4°C. Post-target capture products were cleaned up using 2.2X KAPA Pure Beads (Roche Sequencing Solutions, Indianapolis, IN, USA).

Amplified DNA segments were prepared for sequencing with 1.5 µL of indexed primers and adaptors and 12.5 µL of Q5 High-Fidelity 2X Master Mix (New England Biolabs, USA) in a second-round PCR. The indexing PCR thermocycler program included denaturation at 98°C for 30 s, amplification with 25 cycles at (98°C for 10 s, 65°C for 75 s), final extension at 65°C for 5 min, and hold at 4°C. Post-indexed products were cleaned up with 1.2X beads. Finally, library products were sequenced on the NextSeq 2000 system (Illumina, San Diego, CA, USA) with an average depth of >100,000× per amplicon. Amplicons with coverage less than 10,000× were considered failed.

Sequencing of matched gDNA from white blood cells (WBCs) in samples positive for hotspot mutations was performed using the same amplicon-based sequencing protocol as that used for cell-free DNA (cfDNA) to reduce the confounding effects of mutations linked to CHIP.

### Variant calling, filtering, and annotation

The raw FASTQ data from amplicons were first processed to remove adapters using Trimmomatic (v0.39) (18). The cleaned reads were then aligned to the human reference genome (GRCh38) using BWA-MEM (v0.7.15). Subsequent steps included sorting and marking duplicates with PICARD (v2.25.6) and assessing alignment quality metrics using CollectHsMetrics (Picard). Variant calling was performed with the mpileup function from SAMtools (v1.11) (19).

To determine the limit of detection (LOD), we utilized commercial mutation reference standards Tru-Q1 and Tru-Q0 (Horizon Discovery, Cambridge, UK) and titrated somatic mutations at average variant allele frequencies (VAFs) of 3%, 0.5%, 0.1%, 0.05%, and 0%. These mixtures were fragmented using NEBNext DNA Fragmentase (New England Biolabs) to simulate cfDNA length and then processed through the mPCR workflow as described previously. We compared the observed VAF with the expected VAF for each mutation to LOD of the assay. Moreover, negative cfDNA samples isolated from 570 healthy human plasmas were subjected to the same workflow to establish baseline VAF cutoff values for each hotspot mutation and to eliminate false positives (20). A sample was considered positive for ctDNA if at least one mutation was detected with a VAF ≥ the selected LOD. The mean VAF of a sample was calculated as the average of all positive mutations detected.

### SPOT-MAS assay

The isolated cfDNA samples were analyzed using the previously described SPOT-MAS assay (8). This assay simultaneously evaluates multiple ctDNA signatures, including methylation changes in 450 specific regions, genome-wide methylation patterns, copy number variations, fragment length distributions, and DNA end motifs. The SPOT-MAS workflow is comprised of three primary steps:

**Step 1:** cfDNA isolated from peripheral blood undergoes bisulfite conversion followed by adapter ligation, resulting in a single whole-genome bisulfite library of cfDNA.

**Step 2:** A hybridization reaction is performed on this library to capture the target fraction (450 cancer-specific regions). The remaining whole-genome fraction is recovered by collecting the flow-through and re-hybridizing it with probes targeting the adapter sequences of DNA library. Both the target capture and whole-genome fractions are then sequenced to depths of approximately 52X and 0.55X, respectively, using the DNBSEQ-G400 DNA sequencing system (MGI Tech, Shenzhen, China). Sequencing generated 100-bp paired-end reads with a depth of 20 million reads per fraction. The resulting sequencing data were demultiplexed using bcl2fastq (Illumina, CA, USA) to produce FASTQ files. Quality control of these files was performed using FastQC v. 0.11.9 and MultiQC v. 1.12. Data pre-processing yielded four distinct cfDNA feature sets: target methylation (TM), genome-wide methylation (GWM), fragment length patterns, and end motifs (EM).

**Step 3:** These features were then input into a machine learning algorithm to generate prediction outcomes. The binary classification model, previously detailed (8) provides SPOT-MAS scores, which are used to classify samples as either cancerous or healthy.

### Statistical analysis

The Mann-Whitney U test was used for continuous variables, such as age, while the Chi-squared test was used for categorical variables, such as gender. All statistical analyses were conducted with R (version 2023.12.1+402), utilizing standard data analysis packages and the ggplot2 package for visualization. Confidence intervals were calculated using the Wilson method in R (version 2023.12.1+402).

## RESULTS

### Clinical characteristic of cancer and healthy participants

In this study, plasma samples were collected from a cohort of 255 patients diagnosed with one of the five most common cancer types: breast (n=64), colorectal (n=59), gastric (n=62), liver (n=29), and lung (n=41). Moreover, plasma samples were obtained from 304 healthy individuals as controls. The healthy control group had a median age of 50 years (range: 40-79 years) and included 136 males and 168 females. Cancer patients were significantly older than the control group (p < 0.0001, Mann-Whitney test, **Table 1**). The gender distribution was comparable between the cancer and control groups. All cancer patients were treatment-naïve at the time of blood collection. Of the cancer patients, 14.1% were at stage I, 36.1% were at stage II, and 27.8% were at non-metastatic stage IIIA, while staging information was unavailable for 22.0% of patients. Healthy individuals underwent annual health check-ups, had no history of cancer at the time of sample collection, and were monitored for a period of 12 months to ensure their cancer-free status.

**Table 1.**
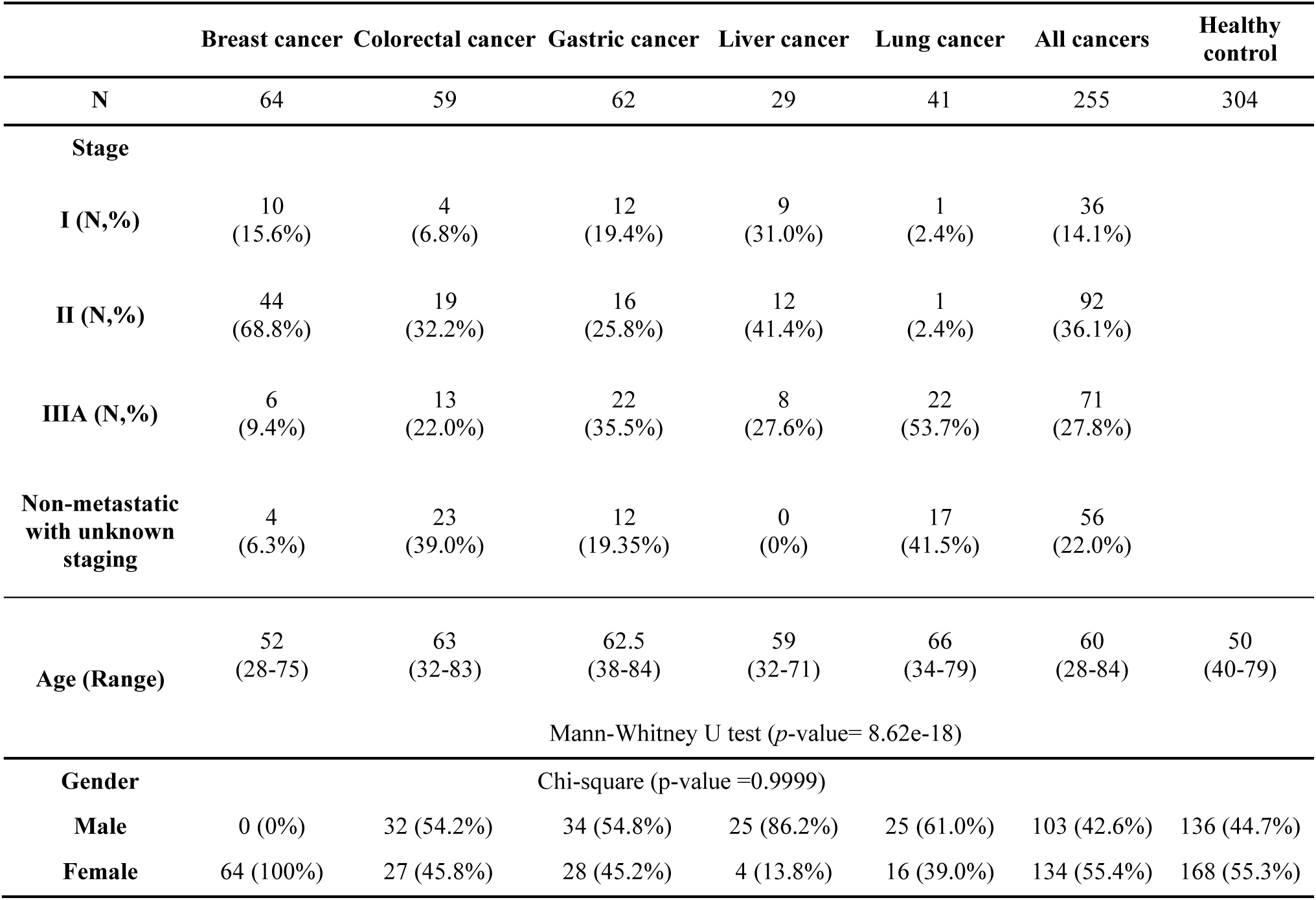
Clinical characteristic of cancer and healthy participants.

|  | Breast cancer | Colorectal cancer | Gastric cancer | Liver cancer | Lung cancer | All cancers | Healthy control |
| --- | --- | --- | --- | --- | --- | --- | --- |
| N | 64 | 59 | 62 | 29 | 41 | 255 | 304 |
| **Stage** |  |  |  |  |  |  |  |
| I (N,%) | 10<br>(15.6%) | 4<br>(6.8%) | 12<br>(19.4%) | 9<br>(31.0%) | 1<br>(2.4%) | 36<br>(14.1%) |  |
| II (N,%) | 44<br>(68.8%) | 19<br>(32.2%) | 16<br>(25.8%) | 12<br>(41.4%) | 1<br>(2.4%) | 92<br>(36.1%) |  |
| IIIA (N,%) | 6<br>(9.4%) | 13<br>(22.0%) | 22<br>(35.5%) | 8<br>(27.6%) | 22<br>(53.7%) | 71<br>(27.8%) |  |
| Non-metastatic with unknown staging | 4<br>(6.3%) | 23<br>(39.0%) | 12<br>(19.35%) | 0<br>(0%) | 17<br>(41.5%) | 56<br>(22.0%) |  |
| Age (Range) | 52<br>(28-75) | 63<br>(32-83) | 62.5<br>(38-84) | 59<br>(32-71) | 66<br>(34-79) | 60<br>(28-84) | 50<br>(40-79) |
| Mann-Whitney U test ($p$-value= 8.62e-18) | | | | | | | |
| **Gender** |  |  | Chi-square (p-value =0.9999) |  |  |  |  |
| Male | 0 (0%) | 32 (54.2%) | 34 (54.8%) | 25 (86.2%) | 25 (61.0%) | 103 (42.6%) | 136 (44.7%) |
| Female | 64 (100%) | 27 (45.8%) | 28 (45.2%) | 4 (13.8%) | 16 (39.0%) | 134 (55.4%) | 168 (55.3%) |

### Profiling hotspot mutations in plasma cfDNA of cancer patients

Recent studies have explored the potential of detecting multiple cancer types through hotspot mutations in plasma cfDNA (13). To assess the effectiveness of this approach, we performed deep amplicon-based sequencing using a panel of 700 hotspot mutations selected from the COSMIC database and somatic mutations in 1,100 Vietnamese cancer patients. Moreover, we performed deep sequencing on matched gDNA extracted from WBCs to exclude mutations associated with clonal hematopoiesis.

Of the 255 cancer patients, 131 (51.4%) exhibited at least one hotspot mutation in plasma cfDNA (**Figure 2A**). The detection rates varied by cancer type, with the highest rate observed in liver cancers (28/29, 92.6%), followed by colorectal cancers (35/59, 59.3%), lung cancers (22/41, 53.7%), gastric cancers (26/62, 41.5%), and breast cancers (20/64, 31.2%).

**Figure 2.**
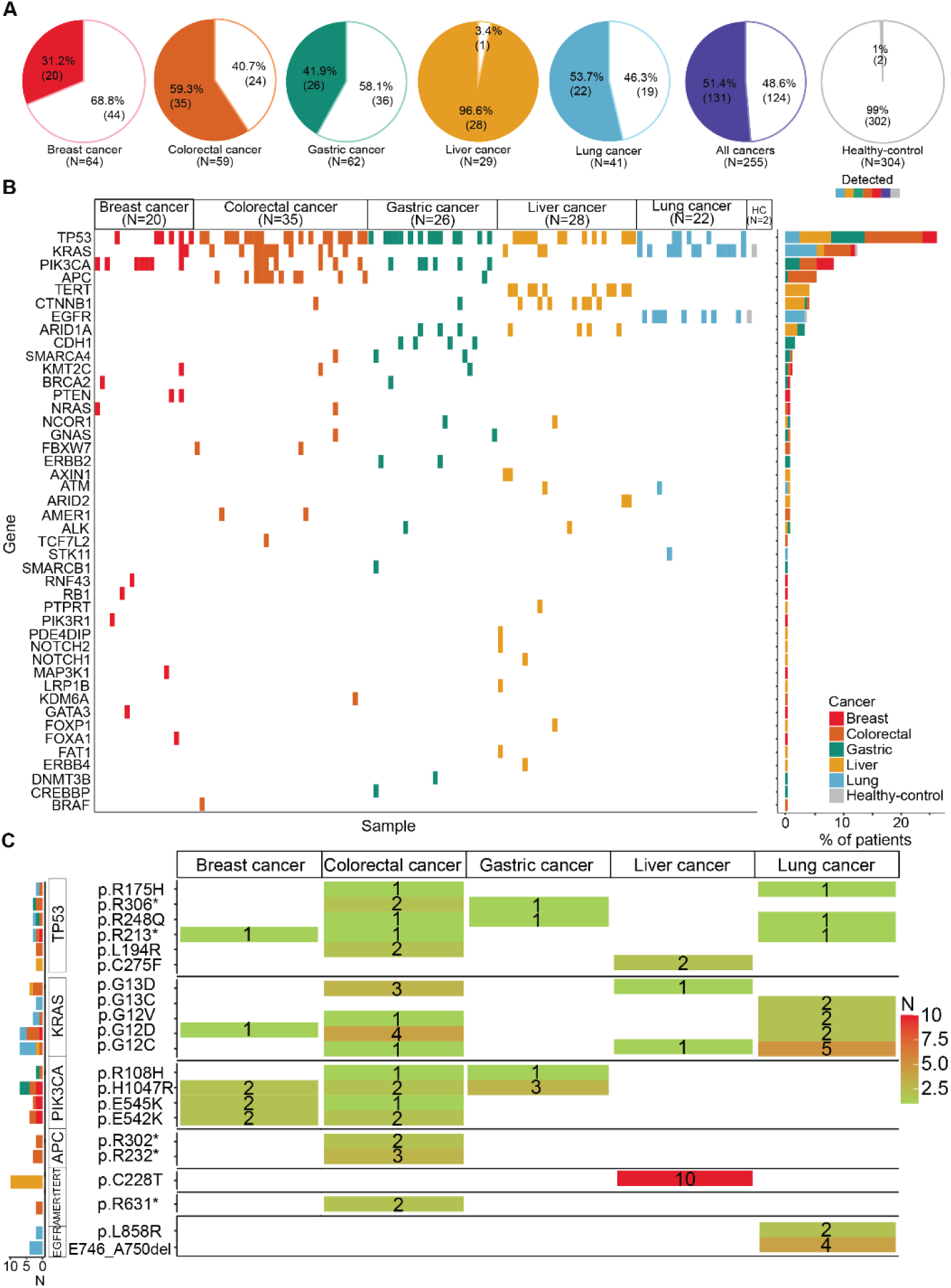
Hotspot Mutation Profiles in cfDNA of Cancer Patients and Healthy Individuals. A. Pie charts illustrating mutation detection rates across five cancer types (breast, colorectal, gastric, liver, and lung). B. Oncoplot showing an overview of mutations in specific genes per cancer type. C. Heatmap illustrating recurrent mutational patterns across and within cancer types

The majority of hotspot mutations originated from *TP53*, detected in 23.1% (59/255) of all cancer patients. *KRAS*-derived mutations were found in 11.0% (28/255) of patients, with the highest prevalence in colorectal (18.6%, 11/59) and lung cancers (29.3%, 12/41). *PIK3CA*-derived mutations were primarily detected in colorectal cancer (11.9%, 7/59), breast cancer (10.9%, 7/64) and gastric cancer (9.7%, 6/62). Notably, *APC*-derived mutations were predominantly observed in colorectal cancer (18.6%, 11/59), while *TERT* promoter and *CTNNB1* mutations were most found in liver cancer, with detection rates of 34.5% (10/29) and 24.1% (7/29), respectively. *EGFR* mutations were exclusively detected in lung cancer patients (19.5%). In the control group of 304 healthy individuals, two were found to carry a hotspot mutation: one with *KRAS* p.Q22K and another with *EGFR* E746_A750del (**Figure 2B**).

We next identified the recurrent mutations (those detected in at least 2 patients) across five cancer types (**Table S3**). For *TP53* mutations, p.R306* and p.R248Q were shared between gastric and colorectal cancers, while p.L194R and p.C275F were repeatedly observed in gastric and liver cancers, respectively (**Figure 2C**). *KRAS* mutations, though most frequently found in colorectal cancer, were also detected in other cancer types, including breast (p.G12D), liver (p.G12C and p.G13D), and lung cancer (p.G13C, p.G12V, p.G12D, and p.G12C). Similarly, *PIK3CA* p.H1047R was shared among colorectal, breast, and gastric cancers. Recurrent *APC* mutations, specifically p.R302* and p.R232*, and *AMER1* (p.R631*) mutations were primarily observed in colorectal cancer. In contrast, *TERT* p.C228T mutations and *EGFR* mutations (p.L858R and E746_A750del) were exclusive to liver and lung cancer, respectively (**Figure 2C**).

Our analysis of hotspot mutations in plasma cfDNA from both cancer patients and healthy individuals showed that 51.4% of patients had at least one hotspot mutation from our panel. Mutations in *TP53, KRAS, PIK3CA, APC*, and the *TERT* promoter were recurrently detected across multiple cancer types or within a single type. These findings highlight the potential utility of our panel in multi-cancer early detection.

### Detection concordance between hotspot mutations and SPOT-MAS

We previously developed a multimodal assay (SPOT-MAS) designed to detect cancer signals in plasma by simultaneously analyzing multiple cancer-specific epigenetic and fragmentomic signatures of cfDNA (8). The assay generates probability scores, known as SPOT-MAS scores, through a machine learning classifier (**Table S4**). A sample is predicted to be cancerous if the SPOT-MAS score exceeds a cut-off value of 0.60. To assess the concordance between cancer detection using hotspot mutations and the SPOT-MAS assay, we set a cut-off for cancer detection by hotspot mutations at a mutant allele fraction (MAF) of 0.05. We observed varying concordance rates across the five cancer types analyzed (**Figures 3A-3D**). Notably, 96.6% of liver cancer patients were detected by both hotspot mutations and the SPOT-MAS assay (**Figure 3D**). However, the concordance was lower for the other cancer types. Breast cancer patients had the lowest concordance rate at 15.6% (**Figure 3A**), followed by gastric cancer (29.7%, **Figure 3C**), colorectal cancer (35.6%, **Figure 3D**), and lung cancer (51%) (**Figure 3E**, blue). Interestingly, hotspot mutations were uniquely detected in 23.7% of colorectal cancer patients, a rate slightly higher than that of the SPOT-MAS assay (23.7% versus 22.0%, **Figure 3B**). In contrast, the SPOT-MAS assay uniquely detected 35.9% of breast cancer patients (**Figure 3A**), 31.2% of gastric cancer patients (**Figure 3B**), and 29.0% of lung cancer patients (**Figure 3E**), surpassing the detection rates observed by the hotspot mutation-based approach (15.6%, 10.9%, 2.0%, respectively). The discordance in detection between hotspot mutations and the SPOT-MAS assay highlights the potential advantage of using hotspot mutations to complement SPOT-MAS to improve detection rates across a broader range of cancer types.

**Figure 3.**
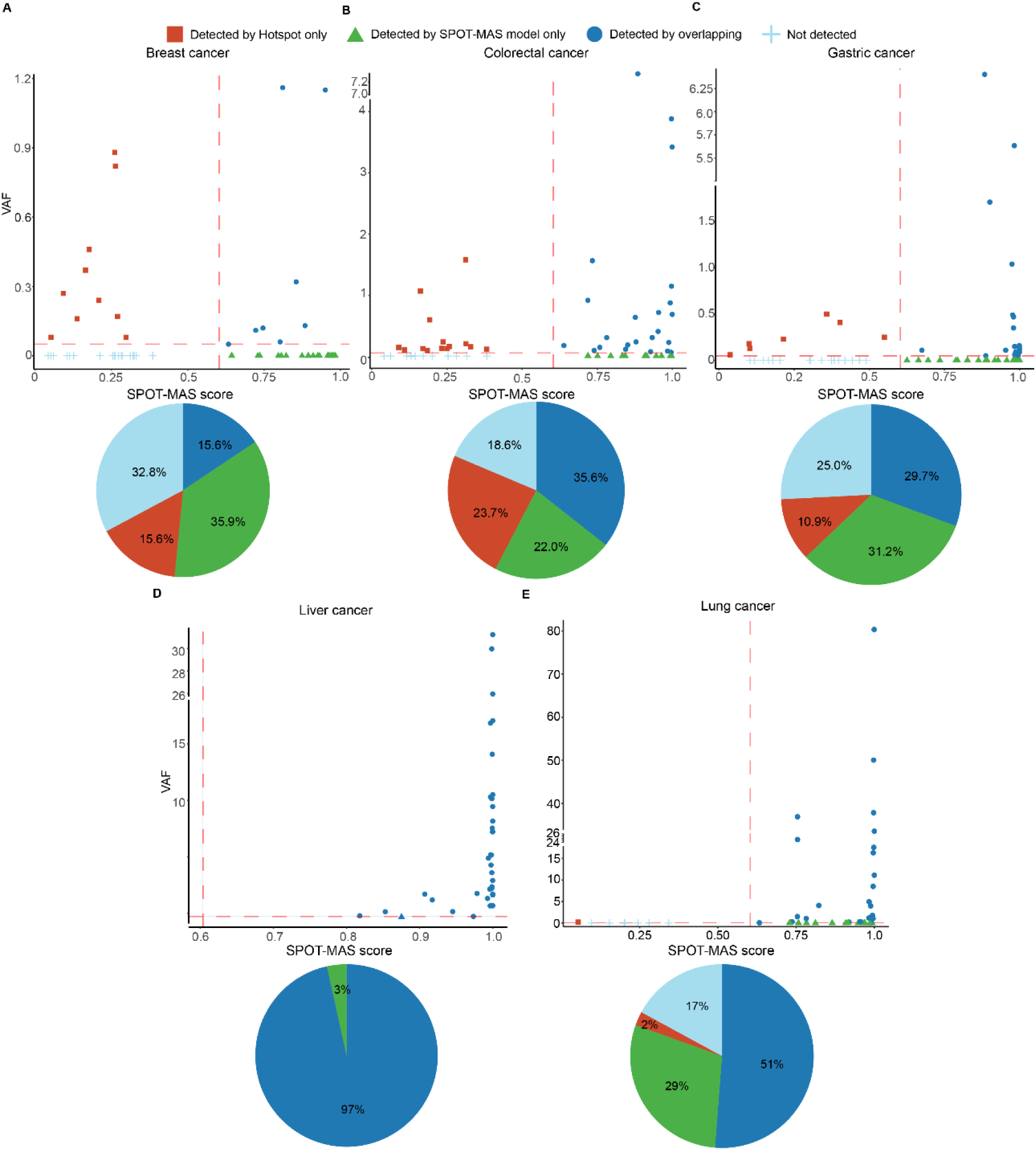
Concordance of Cancer Detection by Hotspot Mutation and SPOT-MAS Assay. A-E. Correlation plots describing MAF and scores from the SPOT-MAS model. The cut-off values were set at 0.05 for MAF and 0.60 for the SPOT-MAS score. Pie charts show the detection rates by hotspot mutations only, SPOT-MAS assay only, both methods, or not detected for breast, colorectal, gastric, liver, and lung cancer patients.

### Integration of hotspot mutations into SPOT-MAS assay enhanced the multi-cancer detection rates

We next explored the potential of combining hotspot mutations with the SPOT-MAS assay to improve detection rates across five cancer types (**Table S5**). Our analysis revealed that SPOT-MAS achieved higher overall sensitivity for detecting cancer patients compared to the hotspot mutation-based approach (65.9% [95% CI: 59.9-71.4] versus 51.4% [95% CI: 45.3-57.4], **Figure 4A**). This advantage was especially pronounced in cancers with a low mutational burden, such as breast cancer (51.6% [95% CI: 39.6-63.4] versus 31.3% [95% CI: 21.2-43.4], **Figure 4A**) and gastric cancer (62.9% [95% CI: 50.5-73.8] versus 41.9% [95% CI: 30.5-54.3], **Figure 4A**). Importantly, both methods demonstrated high specificities, with 99.3% for hotspot mutations and 98.4% for SPOT-MAS. This suggests that combining hotspot mutation data with SPOT-MAS scores using an ‘OR’ rule could enhance detection rates for multiple cancer types while maintaining high specificity. We showed that this combined approach increased overall sensitivity to 78.4% (95% CI: 73.0-83.0, **Figure 4A**) at a specificity of 97.7% (95% CI: 95.3-98.9, **Figure 4A**). The improvement was particularly significant for colorectal, breast, and gastric cancers. When stratifying cancer detection by stages, the hotspot mutation approach had lower sensitivity compared to SPOT-MAS for detecting stage I (44.4% [95% CI: 29.5-60.4] versus 63.9% [95% CI: 47.6-77.5], **Figure 4B**) and stage II cancers (48.9% [95% CI: 39.0-59.0] versus 64.1% [95% CI: 54.0-73.2], **Figure 4B**). Our data demonstrated that the combined approach led to improvements in detecting cancers across all stages.

**Figure 4.**
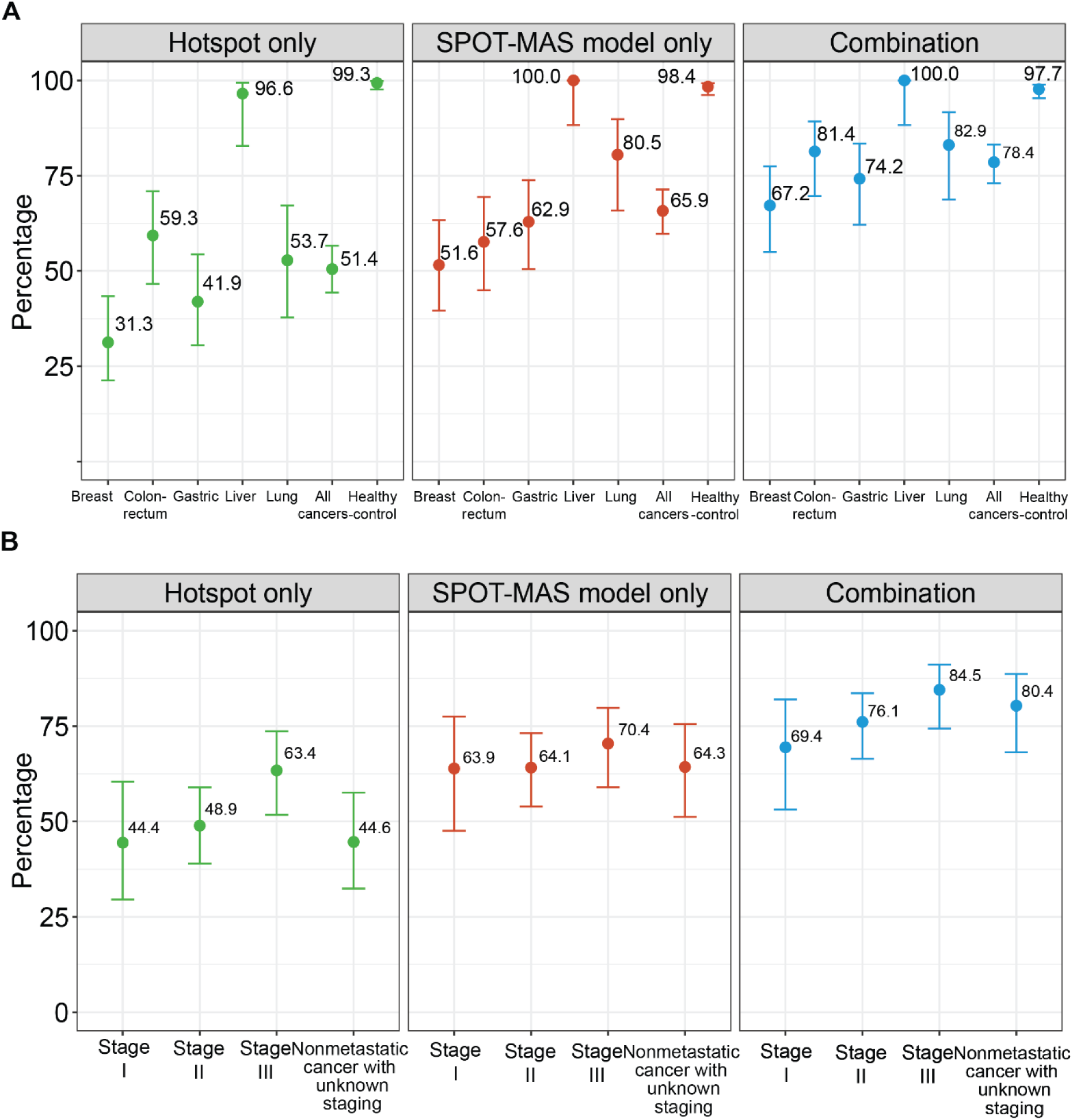
Detection of Cancers Using a Combination Approach. A. Sensitivity and specificity of the hotspot mutation-based approach, SPOT-MAS assay, and combination strategy for differentiating patients with five cancer types from healthy individuals. Data are shown as the mean and 95% confident intervals. B. Bar chart showing the stratification of detection rates by cancer stages.

In summary, our study demonstrated that integrating hotspot mutations into the SPOT-MAS assay could enhance cancer detection across multiple types and stages of cancer.

## DISCUSSION

Liquid biopsy based on multimodal analysis of ctDNA has emerged as a promising approach for MCED (8, 21). Both hotspot mutations and epigenetic features, including methylation and fragmentomics, have been utilized for early cancer detection (22). Our group previously developed an MCED assay, SPOT-MAS, which profiles methylation and fragmentomics features of cfDNA to detect five common cancer types. In this study, we demonstrated that hotspot mutations can complement our SPOT-MAS assay to further improve the sensitivity of cancer detection.

We designed a panel covering 700 hotspot mutations selected from the COSMIC database and somatic mutations identified in Vietnamese cancer patients. This extensive panel identified mutations in 51.4% of cancer patients with a specificity of 99.3%, indicating their potential as biomarkers for multi-cancer detection. We noted that *KRAS* Q22K and *EGFR* E746_A750del were detected in the plasma of 0.7% (2/304) healthy individuals (**Figure 2B**). The detection of hotspot mutations in healthy individuals has been described in previous studies, which suggest that these mutations could derive from very early neoplastic changes or from a normal apoptotic process that eliminates damaged cells with mutations (23).

There was a strong correlation between the detection of hotspot mutations and the SPOT-MAS assay in liver cancer, with 96.6% of liver cancer patients being detected by both methods. This suggests that liver cancer patients may shed high amounts of ctDNA containing both mutations and epigenetic changes. Among liver-associated hotspot mutations, *TERT* promoter mutations were detected in cfDNA from 10 out of 29 liver cancer patients. Notably, hotspot mutations were exclusively detected in 23.7% of colorectal cancer patients who were missed by the SPOT-MAS assay. This finding suggests that colorectal cancer patients may shed ctDNA harboring mutations but not methylation or fragmentomic features detected by SPOT-MAS. This observation aligns with previous studies indicating that colorectal cancers, depending on their subtype or location, exhibit distinct methylation patterns (24). While epigenetic alterations are robust indicators of early tumorigenesis, the variable presence of these signatures in ctDNA underscores the importance of a multimodal approach to improve colorectal cancer diagnosis. On the other hand, SPOT-MAS could exclusively detect 35.9% and 31.2% of breast and gastric cancers, respectively (**Figure 3B and 3C**). Our findings were consistent with previous studies reporting low TMB of these two cancer types (25–27), for which epigenetic changes such as methylation and fragment length demonstrated greater classification power.

Integrating hotspot mutations into the SPOT-MAS assay resulted in enhanced detection sensitivity for colorectal, breast, gastric, and lung cancers (**Figure 4**), highlighting the power of the multimodal approach. Moreover, we observed that SPOT-MAS demonstrated higher detection sensitivity for early-stage cancers (stage I and II) than hotspot mutations, suggesting that methylation and fragmentomic changes are more prevalent in early tumorigenesis (**Figure 4B**) (28).

A limitation of integrating hotspot mutations into the SPOT-MAS assay is the increase in test cost and the requirement for significantly more cfDNA input to accommodate both amplicon and bisulfite sequencing. However, target amplicon sequencing might be the most cost-effective method for detecting hotspot mutations compared to whole exome or whole genome sequencing. Although the five cancer types included in this study are the most common in Vietnamese and Asian populations, it is necessary for future studies to investigate the benefit of combining mutation detection with methylation and fragmentomic profiling in other types of cancers, especially those lacking standard screening. This study is subject to the inherent limitations of a retrospective study and, therefore, requires prospective validation in a larger population.

## CONCLUSION

Integrating hotspot mutation analysis with the SPOT-MAS assay markedly enhances the early detection of multiple cancer types, particularly in the early stages. With an overall sensitivity of 78.4% and specificity of 97.7%, the enhanced assay offers a promising advancement in MCED. Future studies involving larger cohorts are essential to validate these findings and support the broader clinical application of this integrated multimodal approach.

## List of abbreviations

MCED: Multi-cancer early detection
ctDNA: circulating tumor DNA
TNBC: triple-negative breast cancer
NSCLC: non-small cell lung cancer
SPOT-MAS: Screening for the Presence Of Tumor by Methylation And Size
CHIP: clonal hematopoiesis of indeterminate potential
COSMIC: Catalogue of Somatic Mutations in Cancer database
WBCs: White blood cells
LOD: limit of detection
gDNA: genomic DNA
MAF: mutant allele fraction
TMB: tumor mutational burden

## Acknowledgments

We thank all participants who agreed to participate in this study, and all the clinics and hospitals who assisted in patient consultation and sample collection.

## Ethics approval statement

This study was approved by the Ethics Committee of the Medic Medical Center, University of Medicine and Pharmacy and Medical Genetics Institute, Ho Chi Minh City, Vietnam.

## Patient consent statement

Written informed consent was obtained from each participant in accordance with the Declaration of Helsinki.

## Data availability statement

The data that support the findings of this study are available on request from the corresponding author. The data are not publicly available due to privacy or ethical restrictions.

## Tables

**Table S1.** Clinical information of 559 participants

**Table S2.** Details of panel 700 hotspot mutations in 23 genes

**Table S3.** List of recurrent mutations (detected in at least 2 patients)

**Table S4.** Mutation profiling and SPOT-MAS model score of 255 cancer patients

**Table S5.** Detection Accuracy of the combination approach

